# Real world Nuvaxovid COVID-19 vaccine safety profile after 100,000 doses in Australia, 2022-2023

**DOI:** 10.1101/2024.03.17.24304409

**Authors:** Hazel J Clothier, Claire Parker, John H Mallard, Paul Effler, Lauren Bloomfield, Dale Carcione, Jim P Buttery

**Affiliations:** Epi-Informatics, Centre for Health Analytics, Melbourne Children’s Campus, Melbourne, Australia; Epi-Informatics Group and SAEFVIC Epidemiology, Surveillance and Signal Detection, Murdoch Childrens Research Institute, Melbourne, Australia; Department of Paediatrics, University of Melbourne, Victoria, Australia; Melbourne School of Population and Global Health, University of Melbourne, Victoria, Australia; Communicable Disease Control Directorate, Western Australia Department of Health, Perth, Australia; Immunisation Service, Perth Children’s Hospital, Nedlands, Western Australia, Australia; Infectious Diseases Unit, Royal Children’s Hospital Melbourne, Victoria, Australia

**Keywords:** Vaccine safety, adverse events, surveillance, immunisation, COVID, Nuvaxovid, pharmacovigilance, myocarditis

## Abstract

**Introduction:** Nuvaxovid became available in Australia from February 2022, a year later than the first COVID-19 vaccines were released. It was a much-anticipated alternative vaccine for people that had either suffered an adverse event to–and/or were hesitant to receive–one of the mRNA or adenovirus-based COVID-19 vaccines. Although safety from clinical trials was reassuring, small trial population size, relatively low administration rates worldwide and limited post-licensure intelligence meant potential rare adverse events were underinformed.

**Methods:** We conducted a retrospective observational analysis of adverse events following immunisation (AEFI) spontaneously reported to SAFEVAC, the integrated vaccine safety surveillance system used by Victoria and Western Australia, Australia. Reports received from 14 Feb 2022 to 30 June 2023 were analysed by vaccinee demographics, reported reactions and COVID-19 vaccine dose received and compared as reporting rates (RR) per 100,000 doses administered.

**Results:** 356 AEFI reports were received, following 102,946 Nuvaxovid doses administered. Rates were higher post dose 1 than dose 2 (rate ratio 1.5, p=0.0008); primary series than booster (rate ratio 2.4, p<0.0001); in females than males (rate ratio 1.4, p<0.01), especially those aged 30-49 years (RR=1.6, p=0.002).

Serious AEFI included 76 chest pain (RR=73.8), two myocarditis (RR=1.9) and 20 pericarditis (RR=19.4). No cases of Guillain-Barré or thrombosis with thrombocytopaenia syndromes were reported and no deaths attributable to vaccination.

**Conclusion:** The shared SAFEVAC platform enables pooling of clinically reviewed data across jurisdictions, increasing the safety profile evidence-base of novel vaccines like Nuvaxovid and improving the odds for identification and description of rare events across all vaccines.

Key public health message

What did you want to address in this study and why?
Nuvaxovid is a protein-based vaccine for protection against COVID-19. Nuvaxovid received provisional licensure following clinical trials in which 30,058 participants received at least one dose of Nuvaxovid. Introduced following adenoviral vector and mRNA COVID-19 vaccines, some people had been waiting for this vaccine as an alternative to other brands that had risk of blood clots or heart inflammation. We wanted to inform on safety of Nuvaxovid following over 100,000 doses administered in real-world population-wide setting in two Australian states.

What have we learnt from this study?
Adverse events reported were mostly mild, transient symptoms and no deaths were attributed to Nuvaxovid immunisation. Reporting rate for heart inflammation was similar to the mRNA COVID-19 vaccines, but more likely to be milder pericarditis than myocarditis. For over 25,000 people Nuvaxovid was their first dose of a COVID-19 vaccine received, despite being more than a year since COVID-19 vaccines became available.

What are the implications of your findings for public health?
This first real-world population-wide evidence of Nuvaxovid vaccine safety provides reassurance on the risk-benefit of vaccination for protection from severe COVID-19 disease. We must continue to look hard, using real-world data, to not only ensure vaccines are safe, but also support confidence in immunisation.

## Introduction

Nuvaxovid® (NVX-CoV2373, Novavax) is a protein-based COVID-19 vaccine for prevention of coronavirus disease 2019 (COVID-19) caused by SARS-CoV-2, provisionally approved for emergency use by the World Health Organisation since December 2021[1]. While protein sub-unit vaccines have been well established for decades, the Matrix-M™ adjuvant is less familiar, having been used mainly in Ebola vaccines, administered predominantly in low-income countries.

Australia was one of the first countries to use Nuvaxovid in the community, with vaccines administered from mid-February 2022 for primary series in persons aged 18 years and older and extended in July 2022 to individuals from 12 years-of-age and as a booster for those over 18 years[2].

The introduction of Nuvaxovid was almost exactly 12-months following the implementation of adenovirus (Vaxzevria®, AstraZeneca 8 March 2021) and mRNA (Comirnaty®, Pfizer 21 February 2021 and later Spikevax®, Moderna 27 September 2021) vaccines, and over 95% of the eligible population had already received at least one COVID-19 dose[3]. However, Nuvaxovid provided the only available alternate vaccine for people that had either suffered an adverse event to–and/or were unwilling to receive one of the mRNA or adenovirus-based COVID-19 vaccines[4-7]. A US-based survey of 400 individuals hesitant to receive any available COVID-19 vaccines in early 2022 revealed that 55% would likely receive a protein-based vaccine if made available[8].

Provisional registration with the Australian regulatory agency Therapeutic Goods Administration (TGA) was granted on the basis of short-term efficacy and safety data from pooled clinical trial data from approximately 50,000 persons aged 18 years and over, of whom 30,058 received at least one dose of Nuvaxovid, and a paediatric expansion study of 1,487 adolescents (12–17 years of age) administered at least one dose[9-11]. Therefore, the TGA determined “Continued approval depends on the evidence of longer term efficacy and safety from ongoing clinical trials and post-market assessment”[12]. Of particular note were certain adverse events of clinical special interest (AESI) reported infrequently in the clinical trials, but with small numerical imbalances between the Nuvaxovid and placebo arms[13]. These included biliary, neurovascular, and cardiac events (including myocarditis) and uveitis[10, 13]. In addition, one case of Guillain-Barré syndrome was considered as likely associated with the vaccine[14].

Our study provides an epidemiological analysis of Nuvaxovid safety profile, with insights into adverse events following immunisation (AEFI), including frequency, severity and sex distribution following over 100,000 doses of Nuvaxovid administered in a real-world setting.

## Methods

### SAFEVAC surveillance system

SAFEVAC is the integrated vaccine safety surveillance platform used by the states of Victoria (VIC) and Western Australia (WA), Australia[15, 16], covering an estimated 9,163,517 population (VIC 6,503,491, WA 2,660,026)[17].

Reports to SAFEVAC can be made by health care providers, vaccinees (or their guardians) or ascertained from integrated surveillance identifying medically attended potential AEFI reported via active surveillance post-vaccination surveys or linked hospital-immunisation datasets (WA)[18, 19]. Reporting of adverse events is voluntary in Victoria and a statutory requirement for health professionals in WA[20].

Serious AEFI are defined as events resulting in death, are life-threatening, require in-patient hospital admission or prolongation of admission, persistent or significant disability/incapacity, result in a congenital anomaly/birth defect, or require intervention to prevent injury[21].

SAFEVAC accepts all submitted AEFI reports without bias, therefore it is important to acknowledge the adverse events as temporally associated to immunisation and not assessed as causal[16].

### Clinical review

All reports received via SAFEVAC are triaged by an immunisation nurse clinical team, with potential serious AEFI or adverse events of special interest (AESI) triaged for confirmation of the clinical details and follow-up if required. Described symptoms and signs are coded as reaction terms, consistent with medical terminology and/or case definitions as appropriate [15]. As myocarditis and pericarditis were pre-determined AESI for any COVID-19 vaccine, all reports underwent clinical review of reported symptoms, with patient follow-up where contact details were available and consent to contact provided, and categorised according to Brighton Collaboration (BC) case definition level of certainty[22].

### Statistical analysis

Adverse events following Nuvaxovid reported to SAFEVAC from 14 February 2022 to 30 June 2023 were analysed by vaccinee demographics, including reported sex and 10-year age-group; reactions and dose of COVID-19 vaccine received.

Reporting rates were calculated per 100,000 doses recorded for VIC and WA for the same period in the Australian Immunisation Register (AIR) by 28 August 2023[23]. Vaccine dose numbers were referred to as primary series (dose 1 and 2) and boosters (dose 3 or more).

Data were visualised using Microsoft Power BI (Microsoft desktop version 2.118.1063.0, Redmond, WA) and statistical calculations conducted using STATA version 18 (Statacorp, Texas) for 95% confidence intervals (95%CI), rate ratios (RR) with Fisher’s exact test used for group comparisons. A *P*-value less than 0.05 was considered statistically significant. As data on COVID-19 diagnoses were not collected in SAFEVAC, adverse events were not analysed considering any history of prior or coincident COVID-19 infection.

## Results

### Doses administered

A total of 102,946 Nuvaxovid doses were administered during the study period (54,566 VIC, 48,380 WA). Overall, 48.3% of doses were primary series (dose 1 =25,496, dose 2 =24,183, boosters = 53,095), noting this proportion was as high as 74.2% (41,398/55,804) in the first three months of the Nuvaxovid roll-out (Figure 1). Distribution by recorded sex was approximately 20% higher for females (n=56,098) than males (n=46,654) (ratio 1.2). AIR data were missing for age (108), sex (194) and dose (172).

**Figure 1:**
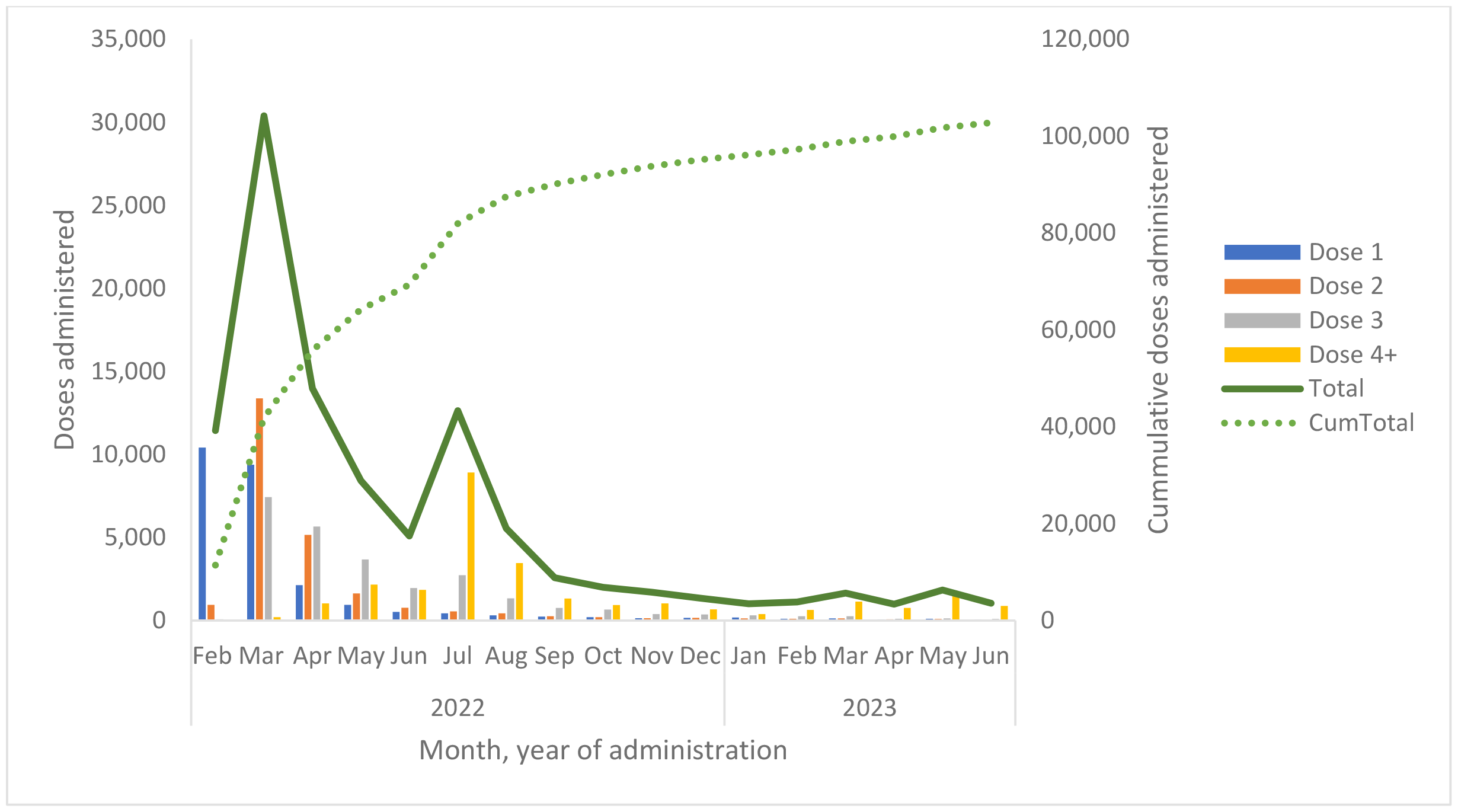
Nuvaxovid doses administered (VIC & WA) by dose and month and cumulative total, 14 February 2022 to 30 June 2023

### AEFI reporting

A total of 356 AEFI reports (143 VIC, 213 WA) were received, being an overall AEFI reporting rate of 345.8 per 100,000 doses. 123 (34.6%) met the definition of serious AEFI (Table 1).

**Table 1:** Characteristics of adverse event reports to SAFEVAC after Nuvaxovid vaccination, WA and VIC, 14 February 2022 to 30 June 2023.

| Nuvaxovid |  | AEFI reports |  |  |  |  |  | Reporter type |  |  | Surveillance system |  |
| --- | --- | --- | --- | --- | --- | --- | --- | --- | --- | --- | --- | --- |
|  | Doses<sup>a</sup> | Total |  | Serious<sup>b</sup> |  | Non-serious |  | HCW<sup>c</sup> | Consumer | Other | n (%) |  |
|  |  | n | Rate<sup>d</sup> | n(%) | rate | n(%) | rate | n(%) | n(%) | n(%) | Passive | Active<sup>e</sup> |
| **Dose 1** | 25,496 | 153 | 600.1 | 54<br>(35.3%) | 211.8 | 99<br>(64.7%) | 388.3 | 68<br>(44.4%) | 80<br>(52.3%) | 5<br>(3.3%) | 126<br>(82.3%) | 27<br>(17.6%) |
| **Dose 2** | 24,183 | 94 | 388.7 | 32<br>(34.0%) | 132.3 | 62<br>(70.2%) | 256.4 | 49<br>(52.1%) | 40<br>(42.6%) | 5<br>(5.3%) | 76<br>(80.1%) | 18<br>(19.1%) |
| **Primary<sup>f</sup>** | 49,679 | 247 | 497.2 | 86<br>(34.8%) | 173.1 | 161<br>(65.2%) | 324.1 | 117<br>(47.4%) | 120<br>(48.6%) | 10<br>(4.0%) | 212<br>(85.8%) | 45<br>(18.2%) |
| **Boosters** | 53,095 | 109 | 205.3 | 37<br>(33.9%) | 69.7 | 72<br>(65.1%) | 135.6 | 45<br>(41.3%) | 36 (33.0%) | 26<br>(23.9%) | 62<br>(56.9%) | 47<br>(43.1%) |
| **Total** | **102,946<sup>a</sup>** | **356** | **345.8** | **123**<br>**(34.6%)** | **119.5** | **233**<br>**(65.4%)** | **226.3** | **162**<br>**(45.50%)** | **156**<br>**(43.80%)** | **38**<br>**(10.70%)** | **264**<br>**(74.1%)** | **92**<br>**(25.8%)** |

<sup>a</sup>Dose not known for 172
<sup>b</sup>Serious AEFI as defined by WHO definition detailed in methods
<sup>c</sup>HCW – health care worker (includes pharmacists)
<sup>d</sup>Rate per 100,000 doses administered
<sup>e</sup>Active – solicited medically attended AEFI or active search of hospital administration data.
<sup>f</sup>Primary series is dose 1 and dose 2 combine

#### Dose

Reporting rate was higher following dose 1 (600.1) than dose 2 (388.7), rate ratio 1.5 (95%CI 1.2, 2.0, p=0.0008) and primary series (497.2) higher than after booster doses (205.3), rate ratio 2.4 (95%CI 1.9, 3.1, p<0.0001). The proportion of reports deemed serious was similar across all doses.

#### Age & sex

AEFI were reported for persons aged from six to 97 years of age [IQR 22] (age unknown for nine cases). Most reports were for females by count (female=219, male=133, neither=1, not stated=3) and rate (rate ratio 1.4, p=0.004 (Table 2)). Reporting rate varied by age-group and sex: higher in females 30–39 and 40–49 years of age (p=0.047 and p=0.021, respectively) and in younger males aged 10–19 years, but without reaching statistical significance (p=0.298) (Figure 2).

**Table 2:** AEFI reactions reported after Nuvaxovid vaccination, as count and rate per 100,000 doses, by sex, presented in order of reporting frequency, VIC and WA, 14 February 2022 to 30 June 2023.

| Reaction (a person may describe >1 reaction per report) | Count<sup>a</sup> |  |  | Rate per 100,000 doses |  |  | F:M rate ratio (p value) |
| --- | --- | --- | --- | --- | --- | --- | --- |
|  | Female | Male | Total<sup>b</sup> | Female | Male | Total |  |
| *Doses administered* | 56,098 | 46,654 | 102,946* |  |  |  |  |
| *Total reports* | 219 | 133 | 356 | 390.4 | 285.1 | 345.8 | **1.4 (0.004)** |
| Chest pain | 47 | 46 | 94 | 83.8 | 98.6 | 91.3 | 0.9 (0.434) |
| Headache | 32 | 26 | 58 | 57.0 | 55.7 | 56.3 | 1.0 (0.934) |
| Fatigue or Lethargy | 33 | 25 | 58 | 58.8 | 53.6 | 56.3 | 1.1 (0.730) |
| Myalgia (muscle pain) | 32 | 14 | 47 | 57.0 | 30.0 | 45.7 | **1.9 (0.041)** |
| ISR<sup>c</sup> (incl pain) | 31 | 13 | 44 | 55.3 | 27.9 | 42.7 | **2.0 (0.034)** |
| Paraesthesia (pins & needles) | 31 | 7 | 38 | 55.3 | 15.0 | 36.9 | **3.7 (0.0006)** |
| Shortness of breath | 21 | 14 | 35 | 37.4 | 30.0 | 34.0 | 1.3 (0.529) |
| Palpitations | 21 | 13 | 34 | 37.4 | 27.9 | 33.0 | 1.3 (0.409) |
| Arthralgia (joint pain) | 17 | 11 | 29 | 30.3 | 23.6 | 28.2 | 1.3 (0.526) |
| Fever | 20 | 6 | 27 | 35.7 | 12.9 | 26.2 | **2.8 (0.021)** |
| Nausea | 22 | 5 | 27 | 39.2 | 10.7 | 26.2 | **3.7 (0.004)** |
| Dizziness | 13 | 13 | 26 | 23.2 | 27.9 | 25.3 | 0.8 (0.642) |
| Myocarditis & pericarditis<sup>d</sup> | 11 | 11 | 22 | 19.6 | 23.6 | 21.4 | 0.8 (0.669) |
| Cardiac symptoms (incl tachycardia) | 5 | 13 | 18 | 8.9 | 27.9 | 17.5 | 0.3 (0.253) |
| Rash | 14 | 4 | 18 | 25.0 | 8.6 | 17.5 | **2.9 (0.049)** |
| Malaise | 12 | 5 | 17 | 21.4 | 10.7 | 16.5 | 2.0 (0.195) |
| Vaccine error<sup>e</sup> | 5 | 11 | 16 | 8.9 | 23.6 | 15.5 | 0.4 (0.068) |
| Abdominal pain | 11 | 2 | 14 | 19.6 | 4.3 | 13.6 | **4.6 (0.030)** |
| Diarrhoea | 9 | 4 | 13 | 16.0 | 8.6 | 12.6 | 1.9 (0.634) |
| Vomiting | 11 | 2 | 13 | 19.6 | 4.3 | 12.6 | **4.6 (0.030)** |
| Lymphadenopathy | 8 | 5 | 13 | 14.3 | 10.7 | 12.6 | 1.3 (0.634) |
| Tachycardia | 5 | 13 | 18 | 8.9 | 27.9 | 17.5 | 0.3 (0.253) |
| Urticaria/hives | 9 | 3 | 12 | 16.0 | 6.4 | 11.7 | 2.5 (0.168) |
| Pruritis | 8 | 2 | 11 | 14.3 | 4.3 | 10.7 | 3.3 (0.117) |
| Influenza-like-illness | 7 | 2 | 9 | 12.5 | 4.3 | 8.7 | 2.9 (0.180) |
| Tinnitus | 7 | 1 | 9 | 12.5 | 2.1 | 8.7 | 5.8 (0.068) |
| Anxiety | 6 | 2 | 8 | 10.7 | 4.3 | 7.8 | 2.5 (0.274) |
| Brain fog | 6 | 2 | 8 | 10.7 | 4.3 | 7.8 | 2.5 (0.274) |
| Anaphylaxis | 7 | 0 | 7 | 12.5 | 0.0 | 6.8 | **6.8 (0.014)** |
| Menstrual changes | 7 | 0 | 7 | 12.5 | 0.0 | 6.8 | **6.8 (0.014)** |
| Pain - other | 4 | 3 | 7 | 7.1 | 6.4 | 6.8 | 1.1 (0.908) |
| Generalised allergic reaction | 4 | 2 | 6 | 7.1 | 4.3 | 5.8 | 1.7 (0.592) |
| Chills | 3 | 3 | 6 | 5.4 | 6.4 | 5.8 | 0.8 (0.829) |
| Angioedema | 4 | 2 | 6 | 7.1 | 4.3 | 5.8 | 1.7 (0.592) |
| Migraine | 4 | 2 | 6 | 7.1 | 4.3 | 5.8 | 1.7 (0.592) |
| Hypertension | 3 | 2 | 5 | 5.4 | 4.3 | 4.9 | 1.3 (0.836) |
| Insomnia | 3 | 2 | 5 | 5.4 | 4.3 | 4.9 | 1.3 (0.836) |
| Altered throat sensation | 3 | 2 | 5 | 5.4 | 4.3 | 4.9 | 1.3 (0.836) |
| Other<sup>a</sup> | 95 | 43 | 138 | 17.0 | 09.2 | 13.4 | **1.8 (0.001)** |
| **Total** | **591** | **336** | **937** | **1053.5** | **720.2** | **910.2** | **1.5 (<0.0001)** |

<sup>a</sup> Reactions with total count <5 not listed to protect privacy
<sup>b</sup> Includes one reaction with sex=neither and for 9 reactions (3 cases) sex was not stated
<sup>c</sup> Injection Site Reaction
<sup>d</sup> Excludes 3 pericarditis and 0 myocarditis cases where an alternate cause was deemed likely .
<sup>e</sup> Vaccine administration errors did not result in an AEFI reaction
\*Total AIR dose count includes 194 for whom sex was not recorded

**Figure 1:**
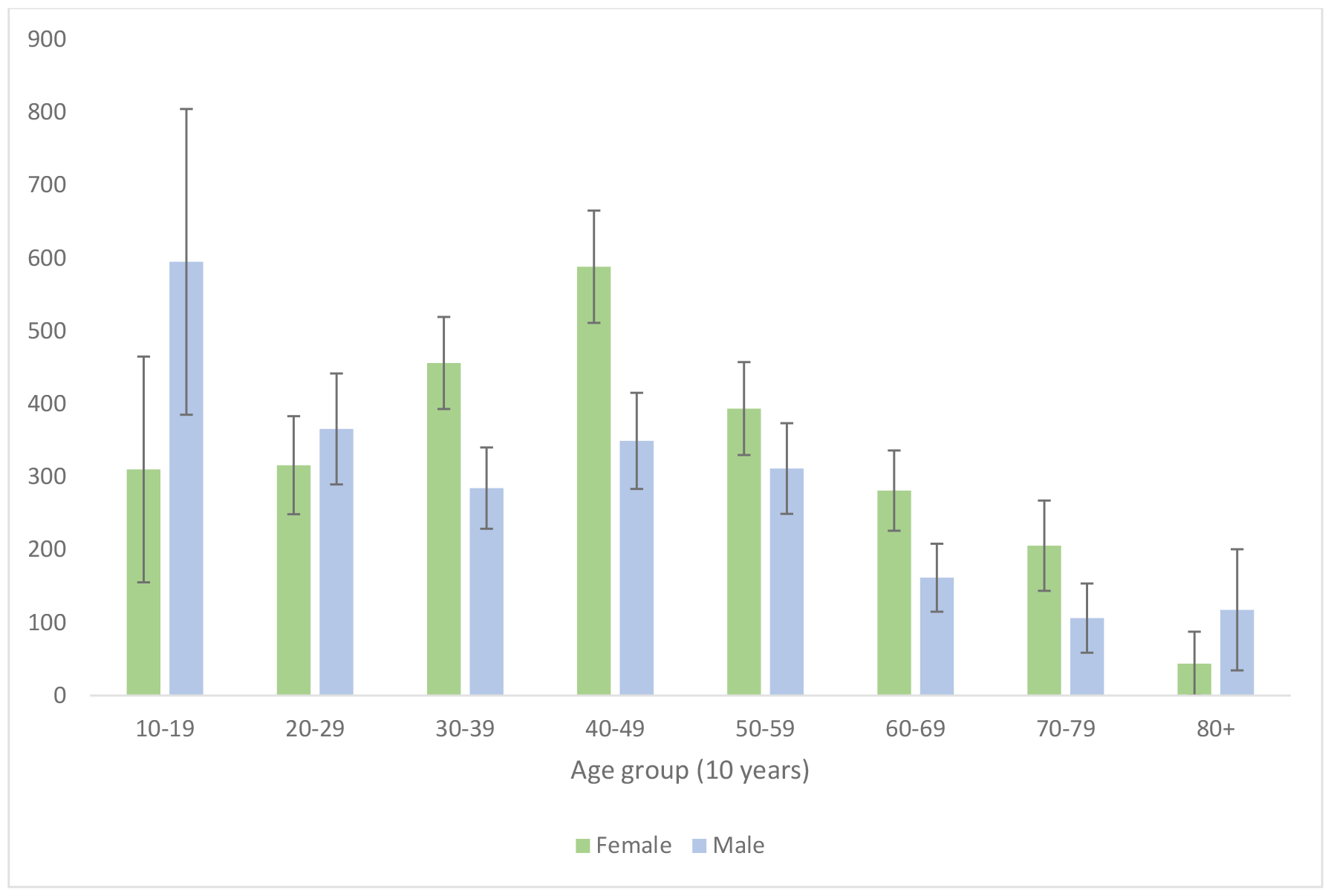
AEFI reporting rate by sex and 10-year age group, with 95% confidence intervals, VIC & WA, Australia *Data for two cases aged <10 years not shown (one male, one female)

#### AEFI reactions reported

From the 356 reports, 937 reactions were described. All reactions reported as total count of five or more are listed in Table 2 by count, sex, rate per 100,000 and female to male reporting rate ratio.

No cases of facial palsy, Guillain-Barré syndrome (GBS) or thrombosis with thrombocytopaenia syndrome (TTS) were reported. Four deaths identified through active search of hospital datasets were identified as occurring temporally proximal to vaccination, however investigation determined no deaths were attributable to vaccination.

Pericarditis was reported post primary and booster doses (total n=20, RR 19.5, 95%CI 11.9, 30.1). Rate was highest following dose 1 (n=10, RR 39.2 95%CI 18.8, 72.1), but similar post dose 2 (n=3, RR 12.4 95%CI 2.6, 36.3) and boosters (n=7, RR 13.2 95%CI 5.3, 27.2). The two clinically confirmed myocarditis cases (BC level 1) were both following dose 2 (overall rate 1.95, 95%CI 0.1, 7.0, or 8.3, 95%CI 1.0, 29.9 specifically for dose 2) with onset within 14 days of vaccination in one male and one female, with no history of myocarditis associated with prior COVID vaccination.

There was one report of uveitis reported on a day 42 active survey response, however insufficient details meant we were unable to follow-up for clinical review and confirmation, including days to onset post the dose 3 Nuvaxovid received.

## Discussion

Our study provides the first southern hemisphere post-authorisation safety profile of Nuvaxovid, describing adverse events reported following over 100,000 doses of Nuvaxovid administered in real-world setting of a high-income country utilizing both spontaneous and active surveillance for AEFI identification.

Commonly reported AEFI were consistent with clinical trial data in that “injection site reactions, fatigue, myalgia, headache, malaise, arthralgia, nausea, or vomiting” all featured in the top 10 reported reactions; and that local and systemic adverse reactions occurred more commonly after dose 2 than after dose 1[10, 11]. However, chest pain was the most commonly reported reaction and shortness of breath and palpitations also featured in the top ten. While the propensity to report chest pain was likely influenced by the acknowledged and broadly communicated risk of myocarditis or pericarditis following mRNA COVID-19 vaccines[24], it was none-the-less a significant incidence of a new associated AEFI with Nuvaxovid and hence chest pain was added to Nuvaxovid product information[11, 25].

We demonstrated slightly higher overall AEFI reporting for females, particularly those aged 30–49 years of age. Sex disproportionality was expected in observed rates of injection site reaction, paresthesia, nausea and vomiting, consistent with pre-licensure studies[26, 27]. However, we also identified disproportional reporting in females for anaphylaxis, which has not been previously reported for Nuvaxovid, although a similar disproportionality had been noted by Somiya *et*.*al[28]* in association with mRNA vaccines. While it was unsurprising that menstrual changes were sex-associated with females, the high reporting rate remained of interest as menstrual changes had not been noted in clinical trials. It is not known if this was because of zero reporting, or because trials were not structured to seek information on menstrual changes[27, 29].

The reporting of heart inflammation post Nuvaxovid was consistent in distribution pattern to that seen with mRNA COVID-19 vaccines, with risk of myocarditis associated with dose 2 but pericarditis more likely to be reported following dose 1 and across any age[30]. The reporting rate for myocarditis following Nuvaxovid was lower than for mRNA vaccines but higher for pericarditis[30]. Our data is not able to distinguish if this was a shift in clinical manifestation or due to a bias in persons seeking to receive Nuvaxovid being more predisposed to—or aware of the need to report—heart inflammation symptoms.

Our findings corroborate the clinical trial results and the single post-licensure study identified from Korea in >18yr olds (14 Feb-31 Dec 2022) with 1,230 AEFI reported via passive and active (text message service on days 0 and 7) following 926,982 doses administered[31]. However, Korea reported an overall lower AEFI reporting rate of 132.7 per 100,000 and lower proportion (7.8%) of serious AEFI. They reported four suspected cases of myocarditis, but no confirmation of cases was described. Korean authors also published a case report of a 30 year-old male with clinically confirmed myocarditis onset 17 days post dose 2 Nuvaxovid as a vaccination complication[32].

In our study, two clinically confirmed cases of myocarditis resulted in a higher overall rate (1.93 per 100,000) than that experienced by Korea of (0.11 per 100,000). Cases were consistent with BC definite level one of certainty but no causality assessment, other than having onset within 14 days of vaccination and no alternate cause noted, was performed.

The shared SAFEVAC platform grants collaborative pooling of clinically reviewed data across jurisdictions, increasing the evidence-base for informing the safety profile of novel vaccines like Nuvaxovid and improving the ability to detect and describe rare events in all vaccines and/or to inform on AEFI in small population groups.

Both states (WA and VIC) clinically reviewed reports potentially indicating an AESI, where medical attention was sought, and all myocarditis and pericarditis reports. This enhancement to routine passive surveillance provides insights to inform diagnosis level of certainty and any subsequent causality assessment, which cannot be informed by reporting alone. Ability to conduct clinical review of reported reactions of myocarditis was important, as the media advocacy related to mRNA vaccine associated myocarditis caused increased propensity to present if experiencing any cardiac symptoms, such as chest pain[33].

This study is based on enhanced passive surveillance data, which will incur the known biases of spontaneous reporting systems, including under-reporting[16]. Although AEFI reporting is not mandated in Victoria this does not result in lower reporting, as is evidenced by Victoria being the lead jurisdiction by AEFI reporting volume nationally[16, 34]. However, in this study AEFI reporting was lower in the larger populated state of Victoria, despite similar number of doses being administered. We hypothesize that pragmatic surveillance modifications made to ensure routine systems were not overwhelmed by the pandemic surge brought intentional focus and increased case ascertainment of medically attended events, such as chest pain[34, 35].

This study aimed to provide a description of AEFI reported following Nuvaxovid and is not structured to provide direct comparison to other brands administered in different phases of circulating COVID-19 disease incidence or surveillance processes initiated to accommodate the high volume of reporting through the pandemic[34, 36]. SAFEVAC collates reports of adverse events and does not have identified data-linkage to disease notification datasets necessary to understand the impact of prior or coincident COVID-19 disease on AEFI occurrence and/or increase in incidence above background rates[37, 38]. Nuvaxovid was the only vaccine introduced in Australia during a period of high circulating COVID-19[36], therefore caution is advised in making direct comparisons of AEFI incidence between brands administered without knowledge and correlation of the inferences made considering different eras of both pandemic and surveillance strategies[39].

Several AEFI showed wide disparity in reporting rate between female and males, notably tinnitus with a rate ratio of 5.8 (p=0.068) and warrant a close watch. A limitation of early analysis or when low numbers of vaccine have been administered is insufficient sample size to inform statistical difference in disproportionate reporting. Collaborative data networks that can pool data or conduct meta-analyses can be beneficial when events are rare or when stratified analyses of less common events is required[37].

Vaccine hesitancy due to innate caution of novel vaccine platforms and fear of blood clots and cardiac inflammation attributed to specific COVID-19 vaccine brands had led to some people “waiting for Novavax” (Nuvaxovid)[4-8]. Our data validates this notion, as despite being introduced nearly 12 months after adenoviral vectored and mRNA COVID-19 vaccines, over 74% of all doses administered in the first three months were as part of a primary series and over 25,000 persons receiving at least one dose of COVID-19 vaccine who otherwise may not have been vaccinated. This indicates not only the importance of enabling choice, but that there is appetite for nuanced vaccine safety profiling.

Our study findings provide broad population-level safety intelligence for Nuvaxovid, however, there is increasing demand toward personalized information or “precision immunisation” and rightly so for true informed consent on individual vaccination risk-benefit. Exploration of linked datasets, or primary care datasets where vaccine administered by usual general practitioner, could assist in ascertaining not only trends in health outcomes but also if the population receiving Nuvaxovid had a differing co-morbidity profile and/or COVID-19 infection status and/or cardiac inflammation with prior COVID-19 vaccination[40] and/or variation in propensity to seek medical attention, for example for cardiac symptoms.

More broadly, further studies are also needed to explore the impact of sex-disproportional AEFI on individual perception of vaccine safety, confidence to vaccinate and whether communication—for example that transient menstrual changes can be anticipated[41]— would allay concerns and avoid inducement of vaccine hesitancy[42, 43].

While our population-wide real-world evidence has bolstered vaccine safety insights, surveillance must continue. Using the statistical estimation rule of three[44], having 100,000 administered doses approximates 95% confidence of detecting rare AEFI with incidence of >1 in 33,333 vaccinations, far less than the 3 million doses required to detect the very rare 1 in a million event, or to detect a doubling increase above expected background rates of a rare events[35]. Therefore, it is imperative to maintain continued vaccine vigilance particularly as we are now seeing multi-heterologous COVID-19 vaccine exposures and new-variant booster vaccine programs are anticipated.

## Conclusion

Nuvaxovid, despite its late arrival into Australia’s COVID-19 vaccination program, played a vital role by providing vaccine choice, increasing uptake with potentially 25,000 additional persons accepting a COVID-19 vaccination dose.

SAFEVAC’s collaborative data model supporting multi-jurisdictional analysis enabled clinical reviewed adverse event information in sufficient volume for meaningful insights. Reassurance that rates of pericarditis and myocarditis presented similarly to the mRNA vaccines overall but skewed toward pericarditis with minimal reports of the more severe myocarditis condition: a finding potentially impacted by channelling of at-risk/hyper-aware persons to receive Nuvaxovid and increased awareness to seek medical attention and investigate chest pain symptoms in the post-vaccination period.

Robust post-licensure vaccine vigilance using real-world data is essential in this era aspiring to global biosecurity with Global Pandemic Preparedness 100 Days Mission[45], to not only ensure safe immunisation, but also to maximise community confidence—as it is not vaccines stored in fridges, but people being vaccinated that saves lives.

## Data Availability

Personal health data are held in confidence by the respective jurisdictional governance bodies and can only be released in accordance with ethics committee approvals. Aggregate summary data are publicly available weekly online in Victoria and annually for both Victoria and Western Australia.

## Ethical statement

Ethical approval for this study was provided in Victoria as use of a registered database 36219 for public health surveillance and by the Chief Health Officer WA for provision of de-identified aggregated data informing public health surveillance. Permission to publish data from the Australian Immunisation Register was granted by Australian Government Department of Health and Aged Care.

## Acknowledgements

We acknowledge Gemma Cadby and Carla Drake-Brockman (WA) for support in data management; Daryl Cheng (VIC) for clinical review of myocarditis and pericarditis cases reported in Victoria and Rebecca Gang support with AIR data (VIC).

## Author contributions

All named authors contributed to the concept and design of this study, had access to data of their respective state/jurisdiction and were involved in the writing, editing and final approval for submission of this manuscript. Hazel Clothier and John Mallard were responsible for analysis of the de-identified data aggregated across both states to provide collated findings.

